# Preserved electrophysiological markers of confidence in schizophrenia spectrum disorder

**DOI:** 10.1101/2022.10.19.22281249

**Authors:** Martin Rouy, Matthieu Roger, Dorian Goueytes, Michael Pereira, Paul Roux, Nathan Faivre

## Abstract

A large number of behavioral studies suggest that confidence judgments are impaired in schizophrenia, motivating the search for neural correlates of an underlying metacognitive impairment. Electrophysiological studies suggested that a specific evoked response potential reflecting performance monitoring, namely the error-related negativity (ERN), was blunted in schizophrenia compared to healthy controls. However, attention has recently been drawn to a potential confound in the study of metacognition, namely that lower first-order performance in schizophrenia compared to healthy controls could lead to a spurious decrease in metacognitive performance among patients. Here, we assessed how this confound might also apply to ERN-blunting in schizophrenia. We used an adaptive staircase procedure to titrate first-order performance on a motion discrimination task in which participants (N = 14 patients and 19 controls) had to report their confidence after each trial while we recorded high density EEG. Interestingly, not only metaperceptual abilities were preserved among patients at the behavioral level, but we also found no electrophysiological evidence for altered EEG markers of performance monitoring. These results bring additional evidence suggesting an unaltered ability to monitor perceptual performance on a trial by trial basis in schizophrenia.

## Introduction

Schizophrenia spectrum disorder (SSD) is a mental condition with severe consequences in terms of cognitive (Gopal and Variend 2005; Schaefer et al. 2013) and social abilities (Lysaker et al. 2013, 2021), and more broadly on quality of life (Davis et al. 2020; Hasson-Ohayon et al. 2015). For two decades, an increasing attention has been drawn to metacognitive abilities in individuals with SSD, with numerous behavioral studies suggesting an impaired ability to calibrate confidence judgments according to performance compared to healthy controls (for reviews, see Hoven et al. 2019; Rouy et al., 2021), paralleled with a substantial number of electrophysiological studies showing performance monitoring impairments in this population (for a review, see Kirschner and Klein 2022). In particular, electrophysiological studies highlighted specific evoked response potentials (ERPs) which are blunted in individuals with SSD, such as the error-related negativity (ERN), the error positivity (Pe), or the feedback negativity (FN) (Kirschner and Klein 2022). Among these ERPs, the blunted ERN is considered the most robust candidate as a trait marker predictive of symptomatology (Kirschner and Klein 2022, for a meta-analysis see Martin et al. 2018). The ERN is a response-locked ERP peaking around 100 ms on frontal midline electrodes following errors (Falkenstein et al. 1990; Gehring et al. 1993) and mostly elicited in choice reaction time tasks (e.g. Flanker task, Simon task) where participants are pressured to respond quickly (typically under 1 s), although an ERN is also found in non speeded tasks (e.g. Rausch et al. 2020). The function reflected by the ERN is not clear, whether it reflects a response conflict or an error-monitoring signaling is still debated (for a review, see Gehring et al., 2012, Ullsperger et al. 2014; Vidal, Burle, and Hasbroucq 2020). It has been shown that ERN amplitude increased with confidence that one made an error (Scheffers and Coles 2000). Boldt and Yeung (2015) have also demonstrated a similar gradation of ERN amplitude as a function of confidence, but their multivariate analysis indicated more robust confidence modulations of Pe amplitude - a subsequent neural marker of error awareness (Murphy et al. 2012).

Considering that individuals with SSD typically underperform in cognitive tasks compared to matched controls, it is important to assess if ERN-blunting simply reflects poorer behavioral performance, or if performance monitoring mechanisms are specifically impaired in SSD. In this respect, Kirschner and Klein (2022) mentioned that among the 21 reviewed studies showing a blunted ERN in individuals with SSD, 12 studies reported comparable performance between groups, while the other 9 studies reported underperformance among patients. The meta-analysis from Martin et al. (2018) revealed a similar pattern of results suggesting that group performance was not predictive of ERN blunting. Yet, it has been shown in healthy participants that task difficulty decreases the amplitude of the ERN (Van der Borght et al. 2016), so we reasoned that fortuitous *comparable* group performance - i.e. a difference in performance between groups that did not reach statistical significance -, might still underlie subtle discrepancies between individuals that are not captured behaviorally, but that might nevertheless contaminate measures such as ERPs. In turn, little can be said about differences in ERN amplitudes between groups that behave similarly on average, but that include individuals with varying degrees of performance. Instead, we argue that performance *matching*, which uses a procedure designed to equate performance between each participant is necessary to assess the specificity of electrophysiological markers of performance monitoring in SSD. In the present preregistered study (https://gitlab.com/nfaivre/meta_scz_public), task performance was controlled via a staircase procedure adapting the amount of sensory evidence (motion coherence) according to individual perceptual abilities. This procedure enabled us to match performance between groups and individuals, and therefore to discuss further whether the typical blunted ERN in individuals with SSD is dependent on task-performance or not.

Here we present the results from EEG data collected in patients and matched healthy controls who performed a visual motion discrimination task. Behavioral analyses of the same cohort of participants revealed preserved metacognitive abilities among individuals with SSD (Faivre et al. 2021). Building upon previous findings showing a blunted ERN in individuals with SSD (Kirschner and Klein 2022; Martin et al. 2018), we conducted EEG analyses: 1) to investigate the occurrence of a blunted ERN in individuals with SSD with matched first-order performance, and 2) to explore whether compensatory neural activity related to confidence (ERN-like or Pe) are found among individual with SSD, which could possibly explain why their metacognitive abilities are preserved (Faivre et al. 2021). Besides matching for performance, we also employed a paradigm which did not enforce speeded responses. By giving participants sufficient time to provide a response, we sought to quantify the electrophysiological correlates of performance monitoring without confounding our results with the detection of “slips” - a category of errors corresponding to incorrect executions of appropriate motor programs (Dehaene, Posner, and Tucker 1994; Reason 1990). Slips typically occur when participants provide a speeded response and immediately realize they pressed the wrong button, a process which differs from evaluating the probability that a decision about noisy sensory information is correct (Desender, Ridderinkhof, and Murphy 2021; Ratcliff and Rouder 1998).

## Methods

Methods and analyses were pre-registered (NCT03140475; https://gitlab.com/nfaivre/meta_scz_public). The study was approved by the ethical committee Sud Méditérannée II (217 R01).

### Participants

Twenty individuals with a schizophrenia spectrum disorder (schizophrenia or schizoaffective disorder, 16 males, 4 females) and 22 healthy participants (15 males, 7 females) from the general population took part in this study. Detailed information about recruitment and data collection are available in Faivre et al., (2020). Five patients were excluded for having excessive artifactual EEG activity (see below), and one for having 208/300 trials with a movement onset < 100ms. Three control participants were excluded based on the following criteria: one because of a non-converging staircase, one with an estimated IQ lower than our inclusion criterion of 70, and one because no EEG data was available. In the end, the EEG analyses presented in this article were conducted on 14 individuals with SSD and 19 healthy controls.

### Neuropsychological and clinical evaluation

Several clinical and neuropsychological evaluations were performed on both participants with SSD and healthy controls. In particular, we assessed premorbid IQ with the french National Adult Reading Test (fNART, Nelson and O’Connell 1978), perceptual reasoning with the WAIS IV - Matrix subtest (Wechsler et al., 2008), depression with the Calgary Depression Scale (CDS, Addington et al. 1992), and cognitive insight with the Beck Cognitive Insight Scale (BCIS, Beck 2004).

### Experimental design

We used a visual discrimination task. Stimuli consisted of 100 moving dots within a circle (3° radius) at the center of the screen. On each trial, participants indicated whether the motion direction of the dots was to the left or to the right by reaching and clicking on one of two choice targets (3° radius circle) at the top corners of the screen with a mouse (Figure 1.A). After 6 seconds without response, a buzz sound rang and a message was displayed inviting the participant to respond quicker. Motion coherence was adapted at the individual level via a 1up/2down staircase procedure in order to match task-performance between groups. Following each perceptual decision, participants were asked to report their confidence about their response using a vertical visual analog scale from 0% (Sure incorrect) to 100% (Sure correct), with 50% confidence meaning “Not sure at all”. (For more details, see Faivre et al. 2021).

**Figure 1.**
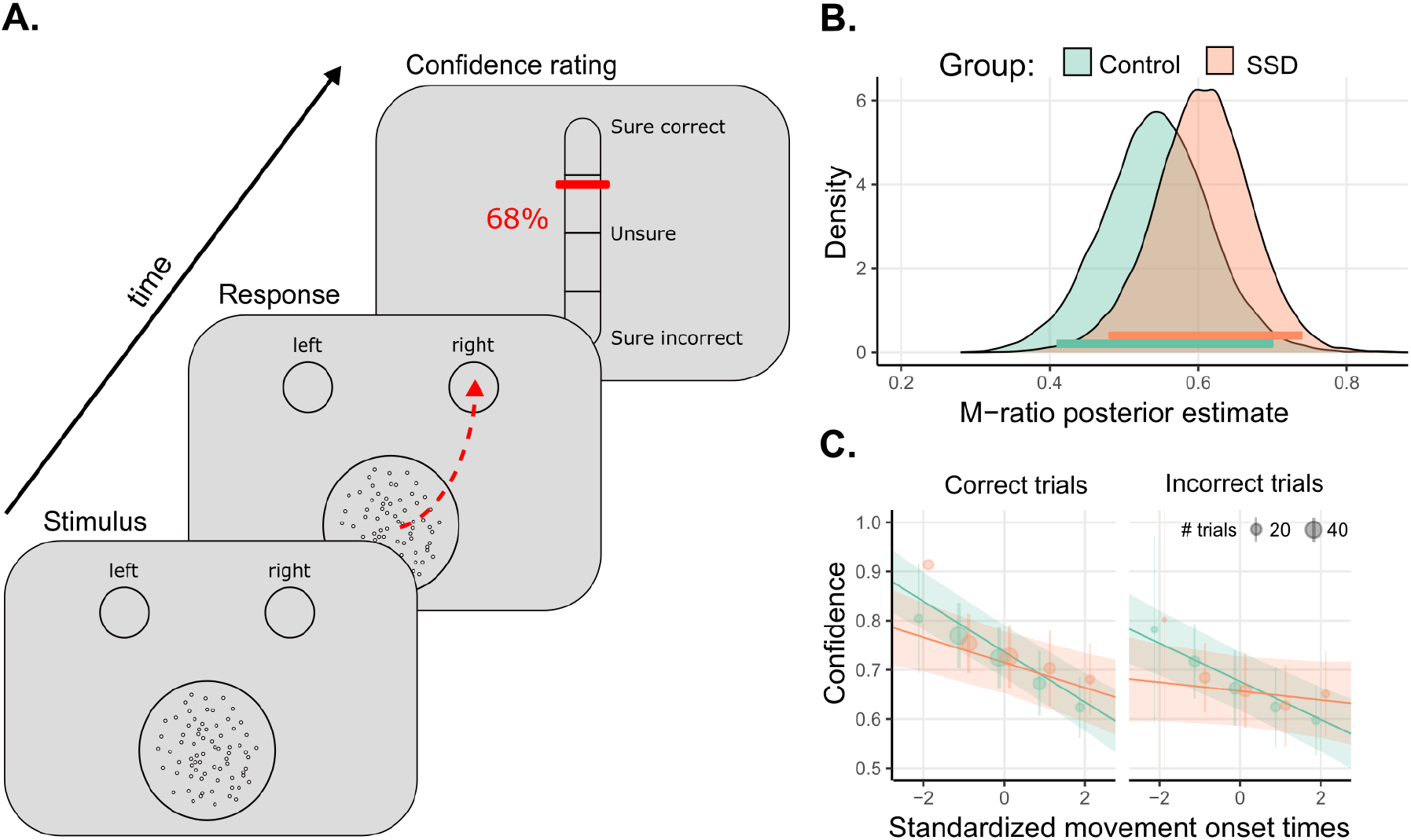
A. Experimental task: Participants indicated the direction of the random-dot kinematogram by clicking in the corresponding response circle, and then estimated their confidence in their response. B. Distribution of posterior estimates of metacognitive efficiency (M-Ratio), horizontal lines depict 95% Highest density intervals. C. Confidence ratings regressed on standardized movement onsets, as a function of groups. Error bars indicate 95% confidence intervals. Orange : Participants with SSD; Green: Control participants. Adapted from Faivre et al. 2021.

In the original study (Faivre et al. 2021), we used mouse trajectories instead of button presses to investigate how the kinematics of mouse movements (velocity and acceleration) related to confidence. Here, we focused on movement initiation rather than response click as a proxy for decision time to avoid the temporal jitter due to kinematic noise in mouse trajectories (i.e. overshoots and undershoots plus small adjustments to reach the response box). We reasoned that time-locking on the initiation of the movement rather than on the response click was of particular relevance when dealing with patients with SSD, who are prone to various motor impairments, either due to medication (Weiden, 2007) or illness (Osborne et al. 2020).. Movement onset was defined as the time needed from stimulus onset to reach 20% of maximum mouse velocity on each trial, from which we subtracted an arbitrary offset of five frames (~83ms) to better capture the moment of movement initiation. Mouse mouvements with velocity peaks lower than 20% of the maximum velocity were discarded as non-decisional, noisy mouse movements. Visual inspection of the corresponding mouse velocity profiles showed that this procedure was effective in finding the movement onset (see Figure S1).

Trials with early (< 100 ms) or late (> 6 s) mouse movements were excluded (6.7 ± 7.1% and 15.3 ± 9.8% of trials in controls and patients, t = -3.06, p < 0.01, BF = 13.2). Changes of mind occurring before the response (i.e., indicated by a change in mouse trajectory, 7.1 ± 5.4% and 8.3 ± 8.1% of trials in controls and patients, resp., t = 0.52, p = 0.61, BF = 0.36) or after the response (i.e., indicated by a confidence lower than 50%, 10.0 ± 12.0% and 10.7 ± 11% of trials in controls and patients, resp. t = -0.17, p = 0.87, BF = 0.33) were excluded, avoiding contamination from additional noise and cognitive processes. We counted as changes of mind trials in which the mouse trajectory - after having reached 20% of the total distance between the initial position of the mouse and the y-projection of the target position - crossed the midline between the two response targets.

### Data analysis

#### Behavioral analysis

Behavioral analyses were performed using R (2020), to ensure that our behavioral conclusions in the original study (Faivre et al. 2021) were still valid on this subset of participants. In particular, we assessed whether our groups of participants were comparable both in terms of demographic and neuropsychological characteristics, and metacognitive performance (i.e. how well participants were able to calibrate their confidence judgments on performance, by computing an index of metacognitive efficiency or M-ratio, (Fleming and Lau 2014) in a Bayesian framework (Fleming 2017) (See pre-registered plan for more details).

#### EEG recording and preprocessing

During the task, EEG activity was continuously recorded using a 64-channelsec HIamp system (g.tec, Schiedlberg, Austria), sampled at 1200 Hz. Electrodes were positioned according to the international 10-10 system with AFz as the reference site. Impedance of electrodes was kept below 5kΩ.

Pre-processing was conducted with Matlab (R2019b) scripts including functions from the EEGLAB toolbox (v2021.0, EEGLAB, Delorme and Makeig 2004). EEG signal was downsampled at 128Hz, then high-pass filtered at 0.5Hz and low-pass filtered at 45Hz. Bad channels were removed manually through visual inspection. Continuous EEG data were locked on mouse movement onset and epoched between -1s pre-movement to 2s post-movement. All channels were re-referenced to the common average, i.e. average over all scalp channels. Horizontal and vertical electro-oculograms were estimated by subtracting AF7 from AF8, and AFz from FPz for later identification and correction of eye-induced artifacts. At this point, noisy epochs with non stereotypical patterns of activity (as opposed to identified non-neural artifactual activity) were excluded through visual inspection. Within each individual data set, the number of components to keep for subsequent independent component analysis (ICA; Delorme, Sejnowski, and Makeig 2007) was obtained through a principal component analysis, keeping only the first components contributing up to 99% of signal variance. ICA was conducted to identify artifactual components before automatic rejection using the EEG artifact detector ADJUST (Mognon et al. 2011). Remaining noisy epochs were excluded by visual inspection. Previously excluded channels were re-interpolated from surrounding channels using spherical splines (Perrin et al. 1989).

To avoid spurious effects of baseline-correction, we only subtracted the average signal per subject and channel over the window from 700 to 200 ms before the movement onset. Finally, to get rid of the remaining noisy trials, we excluded the first and last percentiles of trials by individual, in terms of maximum amplitude.

Among the aforementioned exclusion of five patients for excessive artifactual EEG activity, one patient was excluded because automatic ICA rejection failed to get rid of artifactual components (170/277 trials with regular bursts of voltage amplitude remaining after ICA-based rejection). One patient was excluded because 180 trials and 9 electrodes were rejected after visual inspection. Three patients were excluded for having more than 10 channels with a variance exceeding what was found in the pool of participants by two standard deviations.

#### EEG data analysis

Voltage amplitude was analyzed with linear mixed-effects regressions using R (R core team, 2020) together with the ‘lme4’ (Bates et al. 2014) and ‘lmerTest’ packages (Kuznetsova, Brockhoff, and Christensen 2017). This method allows analyzing single trial data, with no averaging across conditions or participants, and no discretization of confidence ratings (Bagiella, Sloan, and Heitjan 2000). Models were applied to each time sample and electrode for individual trials, to explain broadband EEG amplitude with group and correctness (resp. confidence) as fixed effects, and a random intercept per participant. False discovery rate (FDR) -correction (Benjamini and Yekutieli, 2001) for multiple testing was applied to adjust p-values using the built-in R package ‘stats’. Bayesian mixed-effects regressions with full random-effects structures were fitted using ‘brms’ R package (Bürkner 2017).

Analyses were conducted in three steps: 1) we searched for responsive electrodes, 2) we determined the duration and amplitude of the effect at the level of the cluster of responsive electrodes, and 3) we characterized the robustness of these effects by computing evidence ratios at the cluster level (i.e. the ratio of the evidence supporting the hypothesized direction of the effect, over evidence supporting the non-hypothesized direction of the effect).

1. Search for responsive electrodes: For each time sample, electrode and independent variable of interest (i.e., correctness and confidence), we identified significant effect on voltage amplitude (FDR-corrected) within a time window from 0 to 500ms post-movement onset with the following mixed-effects linear regressions:

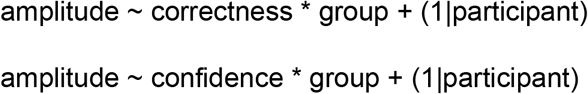 Of note, random slopes were not added at this step as they resulted in convergence failures. The behavioral result of a link between movement onset times and confidence ratings invited us to explore how the ERN-like ERP was modulated by movement onsets with the additional model:

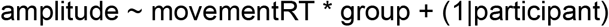

where movementRT refers to movement onset times.
2. Cluster analyses: Building on the literature on ERN, we focused only on central and fronto-central electrodes as regions of interest. To avoid redundant analyses performed on each electrode separately (which are spatially close to each other), and to gain statistical power, we conducted mixed-effects linear regression restrained to the electrodes selected at step 1 which fell within our scalp regions of interest. Regressions were performed at each time sample, taking participants as random intercepts, with electrode nested within participants:

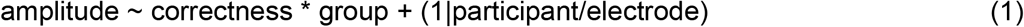

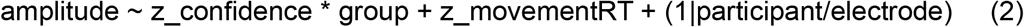

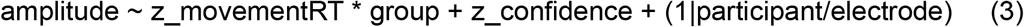

where z_confidence is standardized confidence ratings, and z_movementRT is standardized movement onset times for each participant. Since response times are known to correlate with confidence, z_movementRT was added in model (2), and z_confidence is added in model (3) as covariables of non-interest. Of note, random slopes were not added at this step as they resulted in convergence failures. FDR-correction was applied on the resulting p-values. Only periods with significant adjacent samples extending over more than 50ms were considered genuine effects. The voltage amplitude of the effect of correctness was computed as the average difference between correct and incorrect trials over a 50 ms window centered on the peak of the main effect of correctness. The voltage amplitude of the effect of confidence was computed as the average difference between ‘Very sure’ and ‘Unsure’ tertiles, over a 50 ms window centered on the peak of the main effect of confidence.
3. Bayesian analyses: An evidence ratio was computed on the averaged voltage amplitude over each significant spatio-temporal cluster found in step 2:

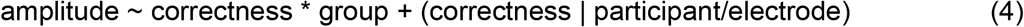

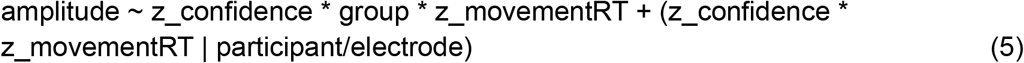 For model (4) we made assumptions leading to the following prior specifications: 1) we assumed that voltage amplitude would be higher for correct versus incorrect responses with a mildly informative Gaussian prior (Mean = 1, SD = 1); 2) we assumed no difference in voltage amplitude between groups with a mildly informative Gaussian prior (Mean = 0, SD = 1); 3) we assumed a blunted ERN among patients with SSD, leading to an interaction effect of group x correctness on voltage amplitude (Gaussian prior Mean = -1, SD = 1) For model (5) we specified the following priors: 1) we assumed that voltage amplitude would be higher (resp. lower) for higher (resp. lower) confidence ratings with a mildly informative Gaussian prior (Mean = 0.5, SD = 1); 2) we assumed no difference of voltage amplitude between groups with a mildly informative prior (Mean = 0, SD = 1); 3) Because we expected a compensatory electrophysiological signal related to confidence among patients with SSD, we assumed an increased confidence ERP among patients with SSD, leading to an interaction effect of group x confidence on voltage amplitude (Gaussian prior Mean = 0.5, SD = 1); 4) We assumed a decreasing amplitude as a function of movement onset times (Mean = -1, SD = 1); and 5) we assumed an interaction effect on amplitude between movement onset times and groups, such that movement onset times from patients with SSD would correlate less with voltage amplitude compared to healthy controls (Mean = 0.5, SD = 1).

Operationalised hypotheses were as follows:

1. The presence of an ERN is indicated by a significant main effect of correctness over frontocentral sites following movement onset,
2. The presence of ERN-blunting is indicated by a correctness x group interaction within the above mentioned cluster of electrodes, characterized by a dampened difference of voltage amplitude between correct and incorrect trials among individuals with SSD compared to healthy controls,
3. The presence of confidence-related compensatory mechanisms in patients is indicated by a confidence x group interaction, characterized by an increased difference of voltage amplitude between confidence levels in patients compared to healthy controls. We did not expect any particular localization or time-window for this effect.

All analysis scripts and behavioral data are publicly available (https://gitlab.com/nfaivre/meta_scz_public). All EEG data will be available upon publication.

## Results

### Demographic and neuropsychological variables

Participants with SSD and healthy controls had similar age, education level and premorbid IQ (see Table 1). However, individuals with SSD were more depressed, with higher levels of cognitive insight than healthy control participants.

**Table 1.**
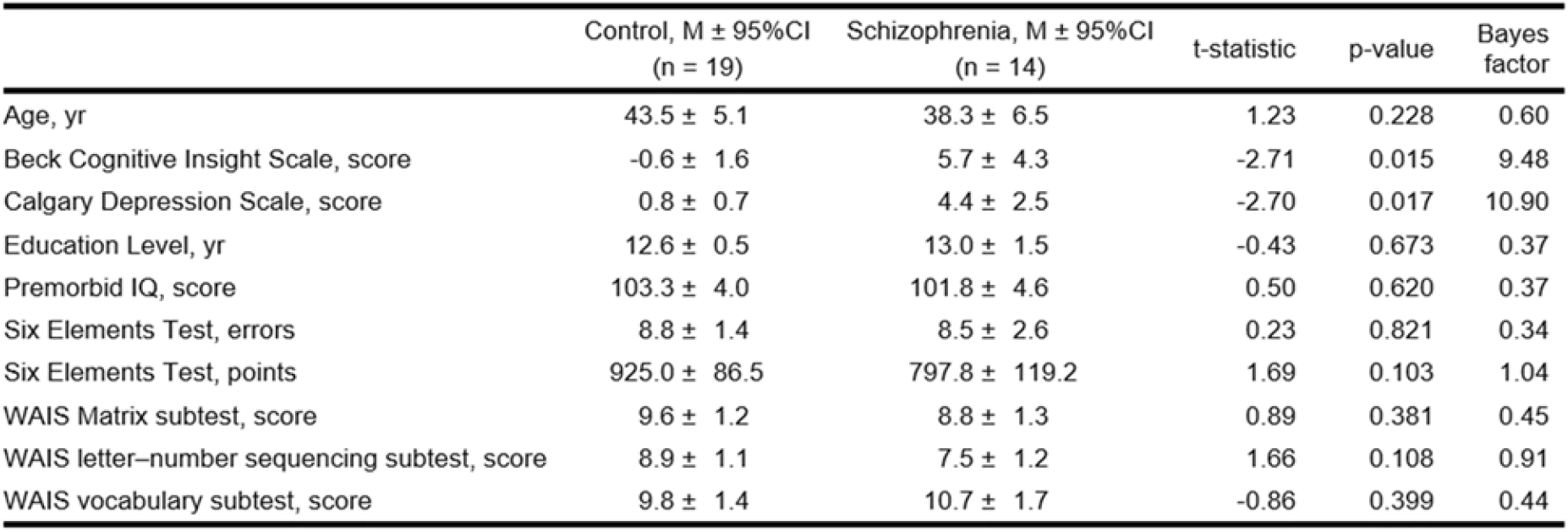
Clinical and neuropsychological characteristics of our sample of participants. CI = Confidence Interval, IQ = Intelligence Quotient; WAIS = Wechsler Adult Intelligence Scale; Bayes Factor > 3 (resp. < 0.33) indicates moderate evidence for H1 (resp. for H0).

| | Control, M ± 95%CI<br>(n = 19) | Schizophrenia, M ± 95%CI<br>(n = 14) | t-statistic | p-value | Bayes<br>factor |
| --- | --- | --- | --- | --- | --- |
| Age, yr | 43.5 ± 5.1 | 38.3 ± 6.5 | 1.23 | 0.228 | 0.60 |
| Beck Cognitive Insight Scale, score | -0.6 ± 1.6 | 5.7 ± 4.3 | -2.71 | 0.015 | 9.48 |
| Calgary Depression Scale, score | 0.8 ± 0.7 | 4.4 ± 2.5 | -2.70 | 0.017 | 10.90 |
| Education Level, yr | 12.6 ± 0.5 | 13.0 ± 1.5 | -0.43 | 0.673 | 0.37 |
| Premorbid IQ, score | 103.3 ± 4.0 | 101.8 ± 4.6 | 0.50 | 0.620 | 0.37 |
| Six Elements Test, errors | 8.8 ± 1.4 | 8.5 ± 2.6 | 0.23 | 0.821 | 0.34 |
| Six Elements Test, points | 925.0 ± 86.5 | 797.8 ± 119.2 | 1.69 | 0.103 | 1.04 |
| WAIS Matrix subtest, score | 9.6 ± 1.2 | 8.8 ± 1.3 | 0.89 | 0.381 | 0.45 |
| WAIS letter-number sequencing subtest, score | 8.9 ± 1.1 | 7.5 ± 1.2 | 1.66 | 0.108 | 0.91 |
| WAIS vocabulary subtest, score | 9.8 ± 1.4 | 10.7 ± 1.7 | -0.86 | 0.399 | 0.44 |

### Behavioral results

Most task-related cognitive variables were comparable between groups. In particular, both groups had similar accuracy levels (SSD: Mean = 0.73, SD = 0.03; controls: Mean = 0.74, SD = 0.02; difference between groups: t = 1.22, p = 0.23, BF = 0.61), which indicated that the staircase procedure was successful in adapting perceptual difficulty (motion variance among SSD: Mean = 1.53, SD = 0.37; controls: Mean = 2.04, SD = 0.40; difference between groups: t = 3.75, p < 0.01, BF = 35.8), with very low dispersion in task performance between participants. There was no difference in average confidence between groups (SSD: Mean = 0.71, SD = 0.12; controls: Mean = 0.71, SD = 0.14; difference between groups: t = 0.0, p = 1, BF = 0.34), indicating no confidence bias, and confidence ratings’ variability was comparable between groups (SSD: Mean = 0.14, SD = 0.05; controls: Mean = 0.16, SD = 0.05; difference between groups: t = 0.72, p = 0.48, BF = 0.41). Furthermore, there was no difference in movement onsets between groups, neither for correct trials (SSD: Mean = 1.23s, SD = 0.72; controls: Mean = 1.32s, SD = 0.66s, difference between groups: t = 0.38, p = 0.71, BF = 0.36) nor for incorrect trials (SSD: Mean = 1.38s, SD = 0.84; controls: Mean = 1.53s, SD = 0.73s, difference between groups: t = 0.53 p = 0.60, BF = 0.38). However, there was a response side bias towards the left in patients with SSD, yet with inconclusive evidence given the BF < 3 (SSD: Mean = -0.32, SD = 0.32; controls: Mean = 0.02, SD = 0.43; difference between groups: t = 2.56, p < 0.05, BF = 2.98).

Concerning metacognitive performance, the Bayesian hierarchical model provided moderate evidence for an absence of difference between the two groups in terms of M-ratio (SSD: Mean = 0.60, 95% highest posterior density interval [95% HDI] = [0.48, 0.74]; controls: Mean = 0.55, 95% HDI = [0.41, 0.70], BF = 0.20), indicating no metacognitive deficits in our sample of participants with SSD (Figure 1.B).

Next, we investigated the relationship between confidence and movement onset. We found a negative relationship between confidence and standardized movement onset (estimate = −0.04 [−0.05 to −0.02]; evidence ratio > 4000), indicating that confidence was higher following earlier movements. This relationship was modulated by the correctness of the responses (interaction correctness x movement onset: estimate = -0.01 [-0.02 to 0.00], evidence ratio = 124) indicating steeper slopes for correct responses, and by the group (interaction group × movement onset: estimate = 0.03 [0.01 to 0.05]; evidence ratio = 67) indicating that confidence ratings were less correlated with movement onset in participants with SSD, compared to healthy controls (Figure 1.C). Yet, the relationship between confidence and movement onset did not interact with correctness x group (interaction correctness x group x movement onset: estimate = 0.00 [-0.02,0.01], evidence ratio = 1.83). Together, these results suggest that in this subsample of patients also, movement onsets are less predictive of confidence than among healthy controls, irrespective of correctness.

### EEG analysis

#### Effect of correctness

Electrodes Cz, C1 and C2 responded significantly to response correctness within the 0-500 ms post-movement window, and were thus selected for further analyses. At the cluster level (Cz, C1, C2), there was a main effect of response-correctness (effect of correctness = -0.34 ± 0.55 μV, evidence ratio = 165) starting 10 ms and until 330 ms after movement onset (Figure 2). The peak of the effect occurred 266 ms after movement onset. However, there was no effect of group, nor a correctness x group interaction effect (evidence ratio = 0.63), providing no evidence for a blunted ERN in patients. The qualitative topographical differences seen in Figure 2 did not reach significance.

**Figure 2.**
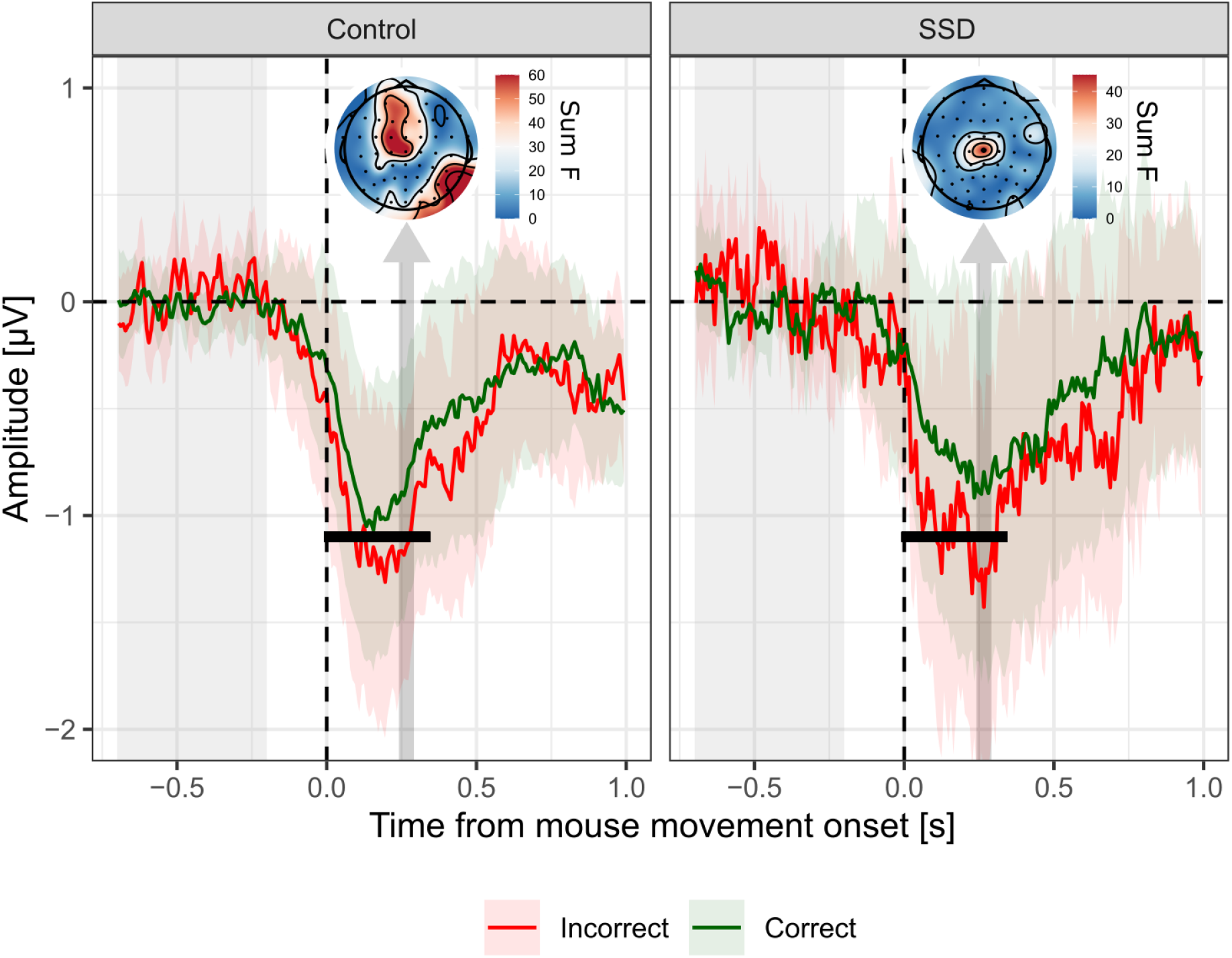
ERP locked on mouse movement onset. Average signal amplitude from central electrodes (Cz, C1, C2) for incorrect responses (red), and correct responses (green). Shaded-areas represent 95% confidence intervals. Light gray shaded area on the left indicates the baseline correction window. Dark gray vertical arrows indicate the 50ms window centered on the peak of the main effect, pointing at the corresponding topographies, scaled according to the magnitude of the main effect of correctness (summed F-values over the 50 ms window). Black horizontal lines indicate adjacent samples with a significant main effect of correctness following FDR correction.

#### Effect of confidence, for correct trials only

To test for a specific effect of confidence irrespective of task performance, we analyzed EEG signals as a function of confidence among correct trials only (thus including 73.7% ± 2.25% of trials in controls and 72.7% ± 2.49% of trials among patients in the analysis, t = -1.10, p = 0.28, BF = 0.54).

Electrodes Cz, C1, C2, FCz, FC1, FC2, FC3, Fz, F1, F2, F3 responded significantly to confidence ratings within the 0-500 ms post-movement window, and were thus selected for further analyses. At the cluster level, there was a main effect of confidence peaking 102 ms after movement onset (effect of confidence = 0.41 ± 0.61 μV, evidence ratio = 12.5) and ranging from - 40 to 227 ms after movement onset (see Figure 3). Importantly, evidence regarding an interaction between z_confidence and group within this cluster was inconclusive (evidence ratio = 1.6). The qualitative topographical differences seen in Figure 3 did not reach significance.

**Figure 3.**
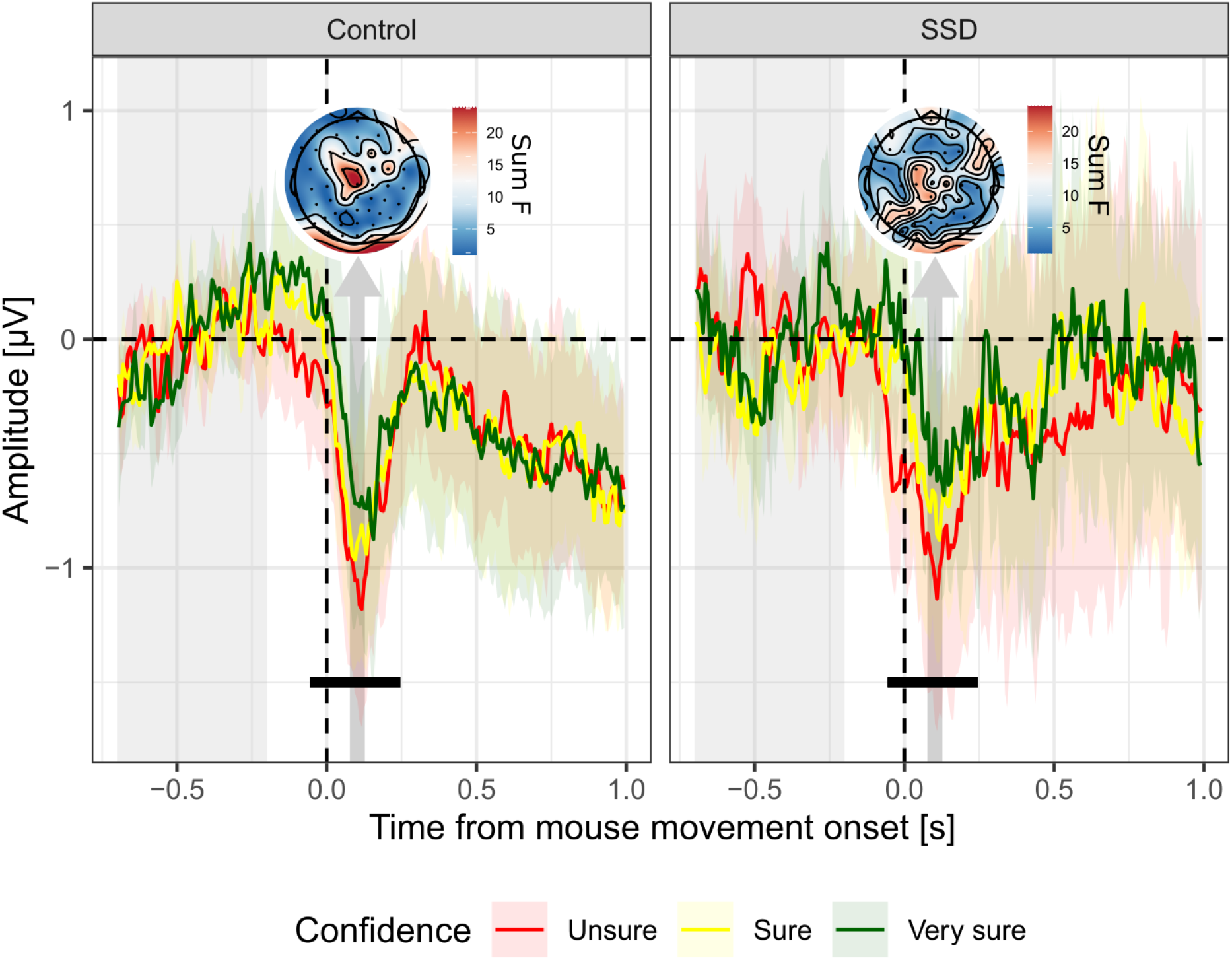
ERP locked on mouse movement onset. Average signal amplitude from fronto-central electrodes (F3, F1, Fz, F2, FC3, FC1, FCz, FC2, C1, Cz, C2) on tertiles of confidence (Unsure, Sure and Very sure trials are plotted in red, yellow and green, respectively). Note that although the graphical representation is based on confidence tertiles, statistical models considered raw continuous confidence ratings. Shaded-areas represent 95% confidence intervals. Light gray shaded area on the left indicates the baseline correction window. Vertical arrows indicate the 50 ms windows centered on the peaks of the main and interaction effects (dark gray and blue, respectively), pointing at the corresponding topography, scaled according to the magnitude of the main and interaction effects of confidence (summed F-values over the 50 ms window). Black lines indicate significant adjacent samples (main and interaction effect of confidence, respectively), following FDR-correction.

Overall, these EEG results are consistent with our previous behavioral results. The comparable ERN-like ERPs observed in both groups (Figure 2) reflect the comparable correctness rates measured behaviorally, and the comparable confidence-related ERPs (Figure 3) reflect the similarity in confidence mean and metacognitive efficiency between groups.

#### Effect of movement onset

As a final analysis step, we sought to investigate the relationship between response times and voltage amplitude, since response times were found to be less correlated with confidence in individuals with SSD at the behavioral level (Figure 1.C). Electrodes Cz, C1, C2, C3, FCz, FC1, FC2, FC3, Fz, F1 responded significantly to response times and were thus selected for further analyses. At the cluster-level, there was a main effect of response times peaking 133 ms after movement onset (effect of movement onset = 0.61 ± 0.95 μV, evidence ratio = 8000) and ranging from -164 to 742 ms after movement onset (Figure 4). Interestingly, an interaction effect between movement onset times and groups was found in two time clusters (depicted with dark blue lines in Figure 4): the first interaction cluster ranged from -39 to 289 ms after movement onset, with moderate evidence (evidence ratio = 5.3). This interaction cluster was characterized by a lesser relationship between EEG amplitude and movement onset times among individuals with SSD (Effect of movement onset = 0.33 ± 0.76 μV, see topography in Figure 4), compared to control participants (Effect of movement onset = 0.67 ± 0.71 μV). The second interaction cluster ranged from 484 to 688 ms after movement onset, with moderate evidence (evidence ratio in favor of the alternative hypothesis = 7.1). This second interaction cluster described the opposite pattern compared to the first one: it was characterized by a steeper increase in voltage as a function of movement onset times among individuals with SSD (Effect of movement onset = 0.38 ± 0.64 μV, see topography in Figure 4), compared to control participants (Effect of movement onset = 0.34 ± 0.74 μV). Furthermore, and in line with the previous analysis on confidence, there was a significant effect of the confidence covariate (depicted as light blue line in Figure 4) from -47 to 133 ms (evidence ratio = 12.5).

**Figure 4.**
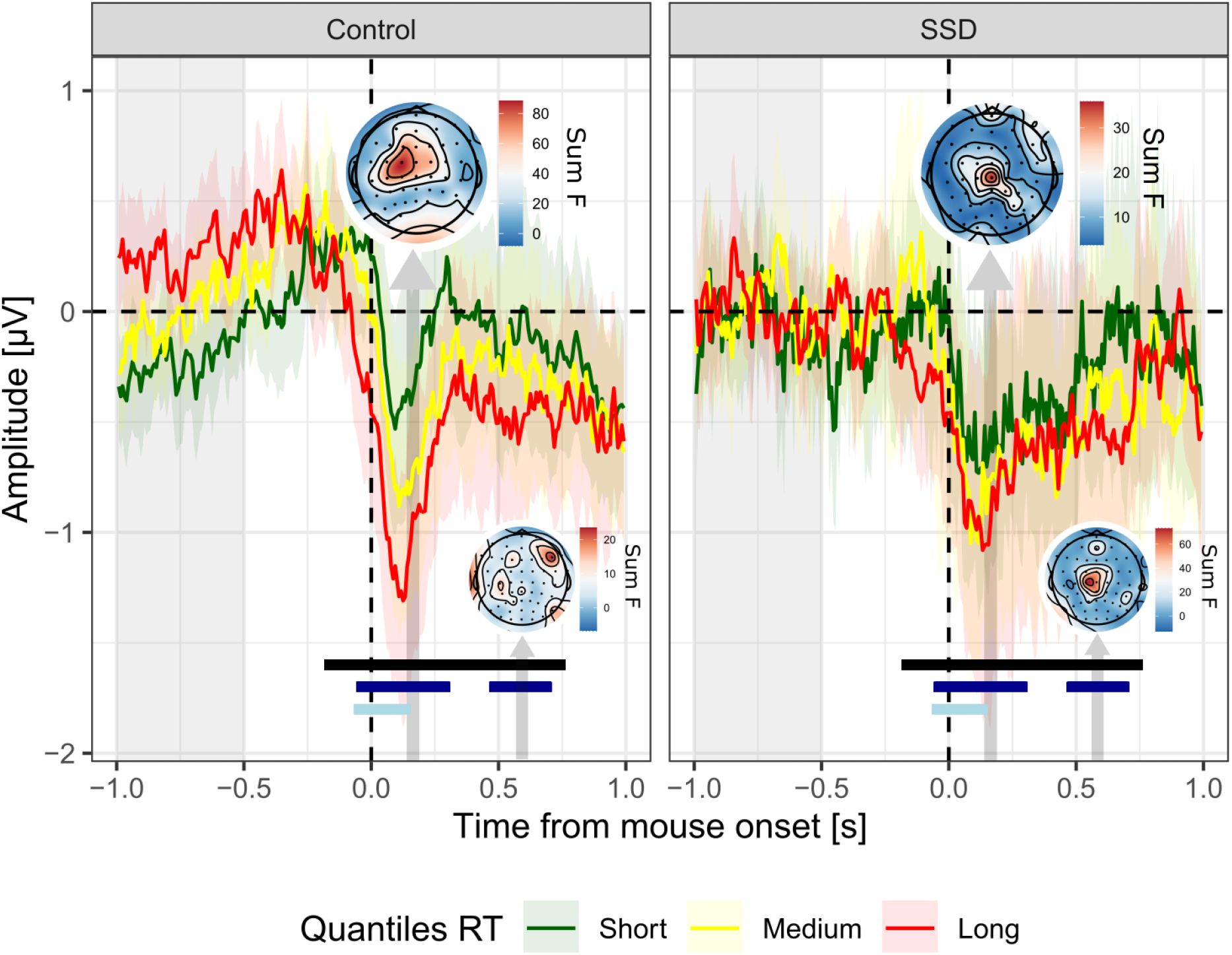
ERP locked on mouse movement onset. Average signal amplitude from frontocentral electrodes (F1, Fz, FC3, FC1, FCz, FC2, C3, C1, Cz, C2) on tertiles of response times (Short, Medium and Long trials are plotted in green, yellow and red, respectively). Note that although the graphical representation is based on tertiles of movement onset times, statistical models considered continuous response times. Shaded-areas represent 95% confidence intervals. Light gray shaded area on the left indicates the baseline correction window. Vertical arrows indicate the 50 ms windows centered on the peaks of the main and interaction effects (dark gray and blue, respectively), pointing at the corresponding topography (N.B.: Occipital electrodes have been deliberately removed from the interaction topography, to focus on the activity of the ROI), scaled according to the magnitude of the main and interaction effects of response times (summed F-values over the 50 ms window). Black, dark blue, and light blue lines indicate significant adjacent samples (main and interaction effect of movement onsets, and main effect of confidence, respectively), following FDR-correction.

## Discussion

In the present study, we sought to investigate EEG data recorded on patients with SSD while they performed a visual discrimination task followed by a confidence rating task (Faivre et al. 2021). Building on the literature on ERN among individuals with SSD (Kirschner and Klein 2022) we expected the ERN-like ERP to be blunted in the group of patients, indicating a performance monitoring deficit under matched levels of task performance between individuals of each group. Then, to make sense of the preserved metacognitive abilities at the behavioral level despite an anticipated performance monitoring deficit among patients with SSD, we hypothesized a distinctive confidence-related ERP among patients, which would constitute evidence for the existence of a compensatory mechanism helping them to provide confidence ratings that are as accurate as those provided by control participants.

We found a negative ERP over frontocentral electrodes that was larger for errors in both groups. Although the peak of this effect occurred later (266 ms) than typical ERN obtained with standard response-conflict tasks, it started 10 ms after the movement onset, consistent with the literature. This difference might be attributed to the fact that we time-locked our analysis onto the initiation of the mouse movement, which might have led to a slightly larger temporal spreading of the ERP. This has the advantage of capturing the very beginning of the decisional process instead of its end-point indicated by a button press (Pereira, Sobolewski, and Millán 2017; Tafuro, Vallesi, and Ambrosini 2020). However, this may be less precise as the definition of a movement onset is temporally more ambiguous than a button press.

In line with our behavioral results, but contrary to our initial hypothesis, EEG analyses revealed no altered neural correlates of confidence among individuals with SSD, which is consistent with the absence of a confidence bias as well as comparable variability in confidence ratings we found behaviorally between the two groups. The fact that evidence in favor of the null hypothesis (i.e., no interaction between confidence and group) remains inconclusive at this stage indicates that future research efforts with more sensitive measures and bigger sample sizes are necessary to conclude if neural correlates of performance monitoring are altered or preserved in SSD. We argue that such research efforts should consider matching performance experimentally between individuals of each group as comparable performance between groups may in itself not be sufficient to disambiguate ERN-blunting from poorer task performance among individuals with SSD. This argument is supported by the finding from Van der Borght et al. (2016) showing that ERN decreases with task-difficulty among healthy participants.

Another reason why no evidence in favor of altered correlates of confidence was found in the present study might be that the ERPs we measured are sensitive to the type of task, i.e. motion discrimination versus response-conflict tasks as commonly employed in the literature (Gehring et al. 2012). Indeed, previous studies on ERN used speeded response-conflict tasks during which participants had to suppress a prepotent response. Here, we were interested in studying errors that arise from the slow accumulation of incorrect noisy sensory information, which are not detected as errors, by giving participants enough time to respond (6s). It might be that the ERN-blunting is specific to fast errors committed in response-conflict tasks, which are known to involve specific mechanisms both in the memory (Ratcliff, 1978) and perceptual domains (Desender et al. 2021; Ratcliff and Rouder 1998).

The difference between fast versus slow errors may be considered as involving “slips” versus “mistakes”, a terminology proposed by Reason (1990). A speeded context increases the proportion of so-called “slips” - a category of errors corresponding to “incorrect executions of appropriate motor programs” (Dehaene et al. 1994) - as opposed to “mistakes”, reflecting inaccurate intentions due to erroneous knowledge. Slips are therefore obvious errors due to participants executing the wrong motor command (pressing A and soon realizing they meant to press B). However, in a non-speeded context, errors are more likely to result from “mistakes” rather than “slips”. In sum, the ERN-blunting in speeded experiments might reflect a specific impairment regarding the monitoring of fast errors or slips, whereas the absence of ERN-blunting in the present non-speeded task might be evidence for a preserved monitoring of genuine mistakes reflected by “slow errors”. Finally, the only notable behavioral difference between the two groups was that confidence was less predicted by response times among patients with SSD (see Figure 1.D). Now extending this aspect to EEG, we found that voltage amplitudes were distinctively modulated by movement onset times among patients, within two spatio-temporal clusters (Figure 4). At first sight, it appears striking that patients with SSD have comparable average response times, confidence levels, metacognitive abilities, and yet lower correlations between confidence ratings and response times compared to healthy controls. Although speculative at this stage, one possibility would be that patients rely more on internal evidence than on additional cues such as response times (e.g. Kiani, Corthell, and Shadlen 2014) or sensorimotor cues (Faivre et al. 2020; Pereira et al. 2020) to rate their confidence.

Of note, our sample of patients with SSD was more depressed than healthy control participants. Interestingly, depression is known to enhance the amplitude of the ERN (for a review, see Bruder et al., 2012), and one might think that it could have compensated for the ERN blunting. Since depression is a very frequent comorbidity in schizophrenia (~ 50%, for a review see Buckley et al. 2009), this confound is most likely present in every ERN study including individuals with SSD, even though it is usually not discussed. Thus, depression is not sufficient to explain the absence of ERN blunting in our sample.

To sum up, we propose two alternative interpretations to explain why we found no evidence for altered neural correlates of performance monitoring among individuals with SSD: 1) Such alterations are confounded with altered task-performance in patients with SSD and are not observed anymore when task-performance is experimentally matched between individuals of each group; 2) Such alterations are specific to “fast errors” committed in response-conflict tasks, which would suggest a specific impairment to detect fast errors or suppress prepotent responses among individuals with SSD.

## Conclusion

In our sample of individuals with SSD showing no metacognitive deficit at the behavioral level, we found inconclusive evidence for deficits in performance-monitoring at the electrophysiological level. Larger scale studies assessing distinct types of errors while finely controlling for task performance are needed to better understand performance monitoring in SSD.

## Acknowledgments

Michael Pereira is supported by a Postdoc.Mobility fellowship from the Swiss National Science Foundation (P400PM_199251). Nathan Faivre has received funding from the European Research Council (ERC) under the European Union’s Horizon 2020 research and innovation programme (grant agreement No 803122). We thank Vincent de Gardelle and Jean-Christophe Vergnaud for their support during data acquisition.

## Contributions

Matthieu Roger, Paul Roux, and Nathan Faivre designed the study and acquired the data. Martin Rouy, Michael Pereira, Dorian Goueytes, and Nathan Faivre analyzed the data. Martin Rouy and Nathan Faivre wrote the article, which all authors reviewed. All authors approved the final version to be published and can certify that no other individuals not listed as authors have made substantial contributions to the paper. This work has been presented at the Association for the Scientific Study of Consciousness in Amsterdam.

## Notes

### Competing Interest Statement

The authors have declared no competing interest.

### Clinical Trial

NCT03140475

### Funding Statement

ERC 803122

### Author Declarations

The study was approved by the ethical committee Sud Mediterannee II (217 R01).

